# Predicting COVID-19 incidence in French hospitals using human contact network analytics

**DOI:** 10.1101/2021.06.17.21258666

**Authors:** Christian Selinger, Marc Choisy, Samuel Alizon

## Abstract

Coronavirus disease (COVID-19) was detected in Wuhan, China in 2019 and spread worldwide within few weeks. The COVID-19 epidemic started to gain traction in France in March 2020. Sub-national hospital admissions and deaths were then recorded daily and served as the main policy indicators. Concurrently, mobile phone positioning data have been curated to determine the frequency of users being colocalized within a given distance. Contrarily to individual tracking data, these can provide a proxy of human contact networks between subnational administrative units. Motivated by numerous studies correlating human mobility data and disease incidence, we developed predictive time series models of hospital incidence between July 2020 and April 2021. Adding human contact network analytics such as clustering coefficients, contact network strength, null links or curvature as regressors, we found that predictions can be improved substantially (more than 50%) both at the national and sub-national for up to two weeks. Our sub-national analysis also revealed the importance of spatial structure, as incidence in colocalized administrative units improved predictions. This original application of network analytics from co-localisation data to epidemic spread opens new perspectives for epidemics forecasting and public health.

**Highlights:**

- We use novel human contact network analytics based on colocation data of mobile app users to follow the dynamics of disease incidence and interventions of COVID-19 in France.
- Time series predictions of hospital incidence are greatly improved by adding these analytics as regressors.
- Sub-national analysis highlights both spatial correlations of incidence and network analytics to obtain high-precision predictions.

## 1. Introduction

The COVID-19 pandemic revealed the importance of identifying early predictors of epidemic dynamics. Indeed, for this infectious disease, hospital admission data is usually the most reliable indicator but it suffers from a two weeks delay with the current state of the epidemic [1, 2]. Screening test results can provide closer monitoring but they often suffer from strong sampling biases.

Mobility data gathered daily by telephone providers and internet services can help to understand epidemic spread, as shown early in the pandemic using individual mobility [3]. More recently, [4, 5, 6] observed that epidemic growth ratios and effective reproduction numbers correlated with human mobility, whereas other studies have used mobility data as early predictors for disease incidence [7, 8, 9]. For instance, [10] used mobile phone location data provided by Google Mobility Trends to calibrate individual movements. However, a limitation of such data is that it only involves individual movement but disease transmission requires at least two people. Here, we use data from Facebook Inc. that informs us on colocation (or colocalization), i.e. not just the position of a single individual but interactions of individuals with respect to their administrative unit of residence (see Methods for details). Similar data has been used by [11] and [12] to simulate meta-population models and investigate the effect of interventions on the resulting network. However, to our knowledge, this data has not yet been used to predict key features of the epidemic such as hospital admissions or deaths.

The body of literature for COVID-19 time series analyses linking epidemiological data to Public Health measures or environmental factors is already widespread, ranging from cross-correlation analysis to complex predictive machine learning tasks [13, 14, 15, 16, 17, 18, 19]. Some methods such as neural networks provide powerful predictions, but the interpretation of the results is offset by the method’s convolutive nature. Here, we choose a middle ground and a more straightforward approach by combining standard Autoregressive Integrated Moving Average (ARIMA) models with original regressors of human contact derived from colocation data.

## 2. Materials and Methods

Facebook Inc. recorded the position of mobile app users, who agreed to turn location tracking on. Every week, users were assigned resident administrative regions based on consistent overnight stays. Given two administrative regions (the ‘départements’) *A* and *B*, we denote by *N*_*A*_(*w*) and *N*_*B*_(*w*) the number of users assigned to each département on week *w*. The colocation probability between *A* and *B* for week *w* is calculated as follows. The week *w* is partitioned into 5 minutes time bins 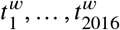. We denote 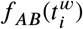 the number of users assigned to *A* and *B* located in the same 600*m*^2^ grid cell within the time bin 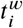 during week *w*. Denoting the total number of colocation events for a given week *w* by 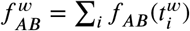, the (symmetric) colocation probability between *A* and *B* for week *w* is then defined by

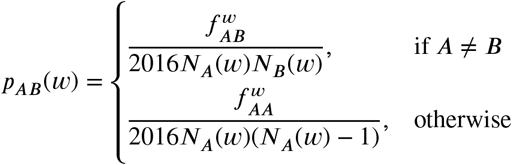

In other words, the colocation probability is the quotient of colocated user pairs over the number of all possible user pairs during a week (with 2016 slots of 5 minutes per week). Importantly, the colocation probability cannot distinguish between events where the same pair of users is colocalised for all 2016 time bins during a week or where a user was colocalized at each time bin with a different user.

For continental France, colocation probabilities were provided at the département level, with a total of *d* = 94 départements covered. Therefore, we constructed 94×94 matrices that capture the weekly colocation probabilities between users depending on their département of residence. These can readily be used to construct weighted contact networks. We also investigated the coverage of Facebook users with assigned département of residence among to French population from the 2019 census data provided by `insee.fr`.

To summarize temporal changes in the resulting weighted contact networks between French départements, we calculated for each week three node-based (clustering, strength and null links) and one edge-based (curvature) graph descriptors. The node-based descriptors were calculated by constructing undirected, weighted graphs from colocation data. For clustering, we used the local clustering coefficient (‘transitivity’ function in the R package igraph), which calculates for a given node all edges weights between the node neighbours relative to the maximal neighbourhood clique size. This clustering coefficient equals one if the node is contained in a clique, *i*.*e*. the node and all its neighbours are connected to each other. The strength of a node is calculated by summing the edge weights incident to the node (‘strength’ function in igraph). The number of null links counts the number of incident edges with zero weight and is bounded from above by *d* −1. A high centrality score for a particular node indicates that colocation between other nodes is decreasing, forcing shortest paths to go through that node, which becomes more central. Curvature was calculated in the sense of discrete Ollivier-Ricci (OR) curvature [20] with a freely available Python code at https://github.com/saibalmars/GraphRicciCurvature. The OR curvature of an edge 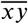 compares the distance between two nodes *x* and *y* with the optimal transport cost W between uniform measures on unit balls centered in *x* and *y*:

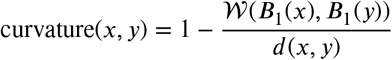

Numerical studies showed that edges with positive curvature tend to be part of a cluster, whereas edges with negative curvature tend to act as bridges between clusters [21, 22, 23]. Therefore, a decrease in curvature can be seen as an indicator for connectivity breakdown.

**Table 1.** Regressors name, description, and source

| name | description | source |
| --- | --- | --- |
| null links | colocation graph | Facebook Inc. |
| strength | colocation graph | Facebook Inc. |
| betweenness centrality | colocation graph | Facebook Inc. |
| clustering | colocation graph | Facebook Inc. |
| curvature | colocation graph | Facebook Inc. |
| between département colocation | colocation data | Facebook Inc. |
| incidence in colocation graph neighbourhood | colocation and incidence data | Facebook Inc. |
| within département colocation | colocation data | Facebook Inc. |
| facebook coverage | colocation data, census data | Facebook Inc., Insee |
| retail and recreation percent change from baseline | average weekly change | Google Mobility Report |
| grocery and pharmacy percent change from baseline | average weekly change | Google Mobility Report |
| parks percent change from baseline | average weekly change | Google Mobility Report |
| transit stations percent change from baseline | average weekly change | Google Mobility Report |
| workplaces percent change from baseline | average weekly change | Google Mobility Report |
| residential percent change from baseline | average weekly change | Google Mobility Report |
| temperature | weekly temperature by département | data.gouv.fr |
| positive test rate | average weekly positive test rate | ourworldindata.org |

For comparison, we also incorporated Google Mobility Report data www.google.com/covid19/mobility/, from which we obtain the daily change in percentage with respect to a pre-pandemic baseline in visits to grocery stores, parks, workplace, residential areas, transit and retail stores. We calculated cumulative weekly changes matching the dates of the weekly recorded colocation data. Since Google Mobility Reports were not available at the département-level, we used these data only for national-level analyses.

We downloaded the positive testing rate, i.e. the ratio of positive tests over all tests from www.ourworldindata.org. To remove reporting bias, we calculated the rolling average with a right-aligned seven-day window and calculated the weekly positive test rate matching the dates of the weekly recorded colocation data.

We downloaded daily temperature data by département which was aggregated into weekly minimum, maximum and average temperature. For the same data, we also calculated quantiles for national analysis.

COVID-19 hospital data for France was downloaded from www.data.gouv.fr. The data comprised daily hospital admissions, ICU admissions, and deaths in hospitals by département. To match the Facebook colocation data, we only considered départements in continental France spanning the time period from March 24, 2020 to March 30, 2021. For the country-wide analysis, we summed the daily incidence in all départements and calculated right-aligned seven-day rolling averages to remove reporting bias. Finally, we calculated cumulative weekly incidence matching the dates of the weekly recorded colocation data. The weekly incidence data were log-transformed prior to the time series analysis.

The time series analysis was performed using the R package forecast. For each week, we trained and tuned Autoregressive Integrated Moving Average (ARIMA) models on historic data starting from March 24, 2020 and performed forecasting for one and two-week horizons. Model parsimony during tuning was determined by the Akaike Information Criterion (AIC). The first four weeks were used for training only, and we started prediction at the fifth week of our records. At any particular week, starting from June 2020, the prediction accuracy was evaluated in terms of mean average error (MAE) between incidence data and predictions for the following two weeks. More precisely, given log-transformed incidence data *y* from week *t*, with exogenous regressors *x*^*i*^, the regression model with ARIMA error [24] is defined by

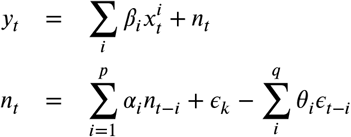

where the regression error *n*_*t*_ is an ARIMA error with *p* the order of the autoregressive part and *q* is the order of the moving average part and *ϵ*_*k*_ are Gaussian errors. Note that *y* and *x* can undergo differencing up to order *d* to ensure stationarity.

We used exogenous regressors based on quantiles of network descriptors, Facebook colocation data, Google mobility reports, and positive testing rates to tune parameters *p, d*, and *q* and to train each model. We then compared the prediction results from models using regressor data to those using historic incidence data alone.

The predictive power of combinations of regressors was assessed by following a step-wise method. For each week, we started using a single regressor and incremented the set of regressors as long as the mean average error of the prediction decreased, thereby yielding a linear combination of regressors with minimal prediction error per week.

For the département-level models, we also applied regression models with ARIMA error. Node-based regressors (e.g. node strength, null links) were based on the actual values, whereas the edge-based curvature regressors consisted of quantiles relative to the département of interest. In addition, for each département, we also used incidence data for hospital admission and mortality in départements with the ten strongest colocation links to the département of interest (denoted by incid_1, incid_2, etc.).

## 3. Results

### 3.1. Network descriptors

The analysis of network descriptors showed a highly dynamic range for the clustering coefficients in relationship with the hospital incidence data and underlying non-pharmaceutical interventions (Fig 1). For instance, in the midst of the first national lockdown in April 2020, the minimum clustering coefficient was at 0.6 but by July 2020 it had regained almost maximum levels (close to 1). The number of null links, which offers a more discrete measure of decreasing connections between départements, reached 72 out of 93 possible null links (77%) for several départements in April 2020. In contrast, several départements exhibited small changes in null links, suggesting that the mobility restrictions impacted the colocation probabilities unevenly across the country. The network strength more than tripled during the same time period in the départements, reaching the strongest colocation probability; a high level which was maintained throughout October 2020, while hospitalization was increasing again in September. The curvature trends followed that of the network strength but were more sensitive to perturbations, for instance around January 1st 2021.

**Figure 1:**
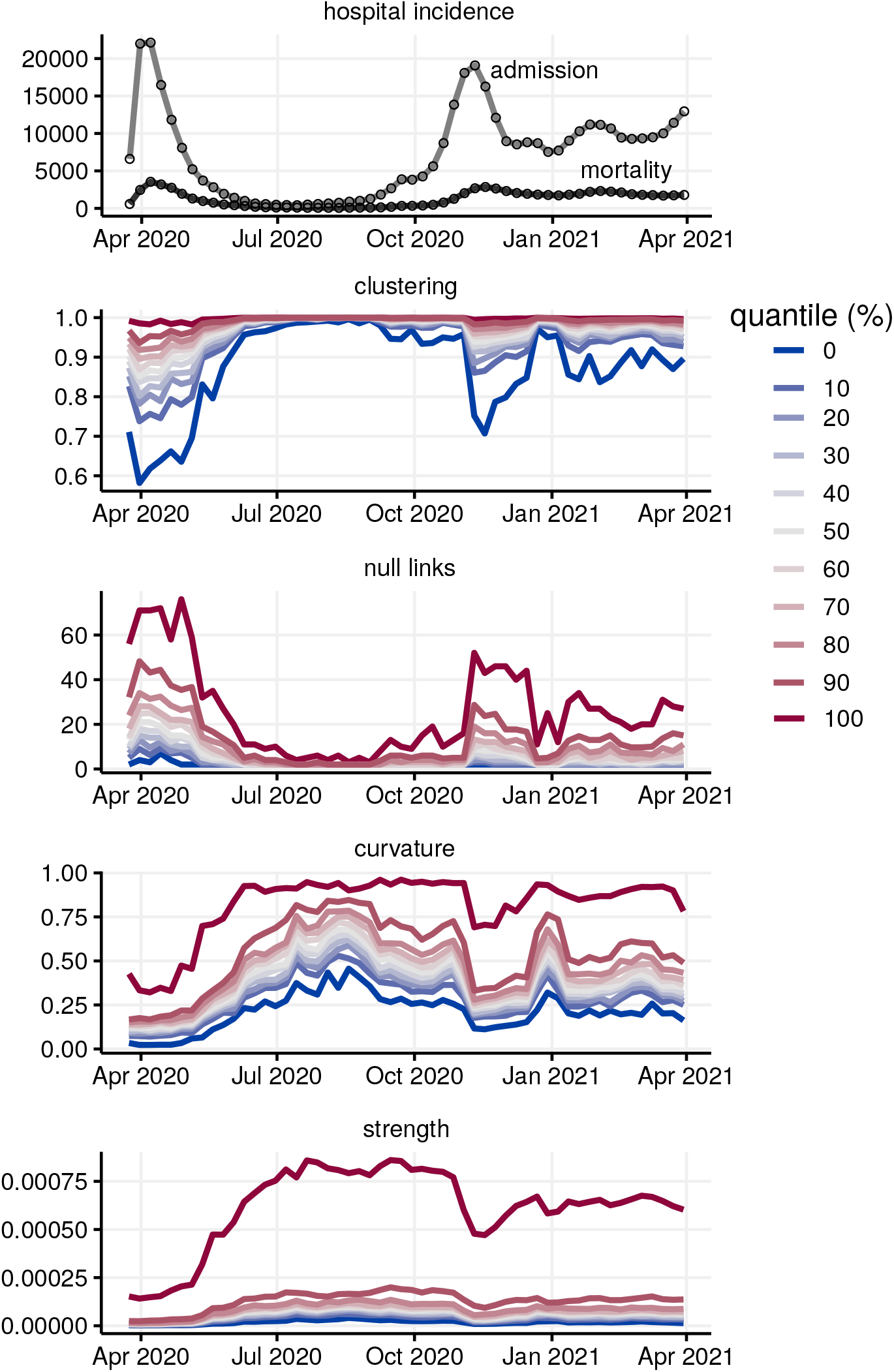
Disease incidence data and network metrics quantiles in France, aggregated by week.

We also investigated spatial snapshots of the network descriptors prior to and during the second lockdown period at the beginning of November 2020. Figure 2 shows, for these two weeks, the 50 strongest colocation weights per week between two départements (red edges) and the number of null links in the départements involved at the end of these links. These result point towards spatial clusters of départements which maintained high colocation weights between each other (*e*.*g*. in the Paris region or in Eastern France) while shutting down connections to many other départements.

**Figure 2:**
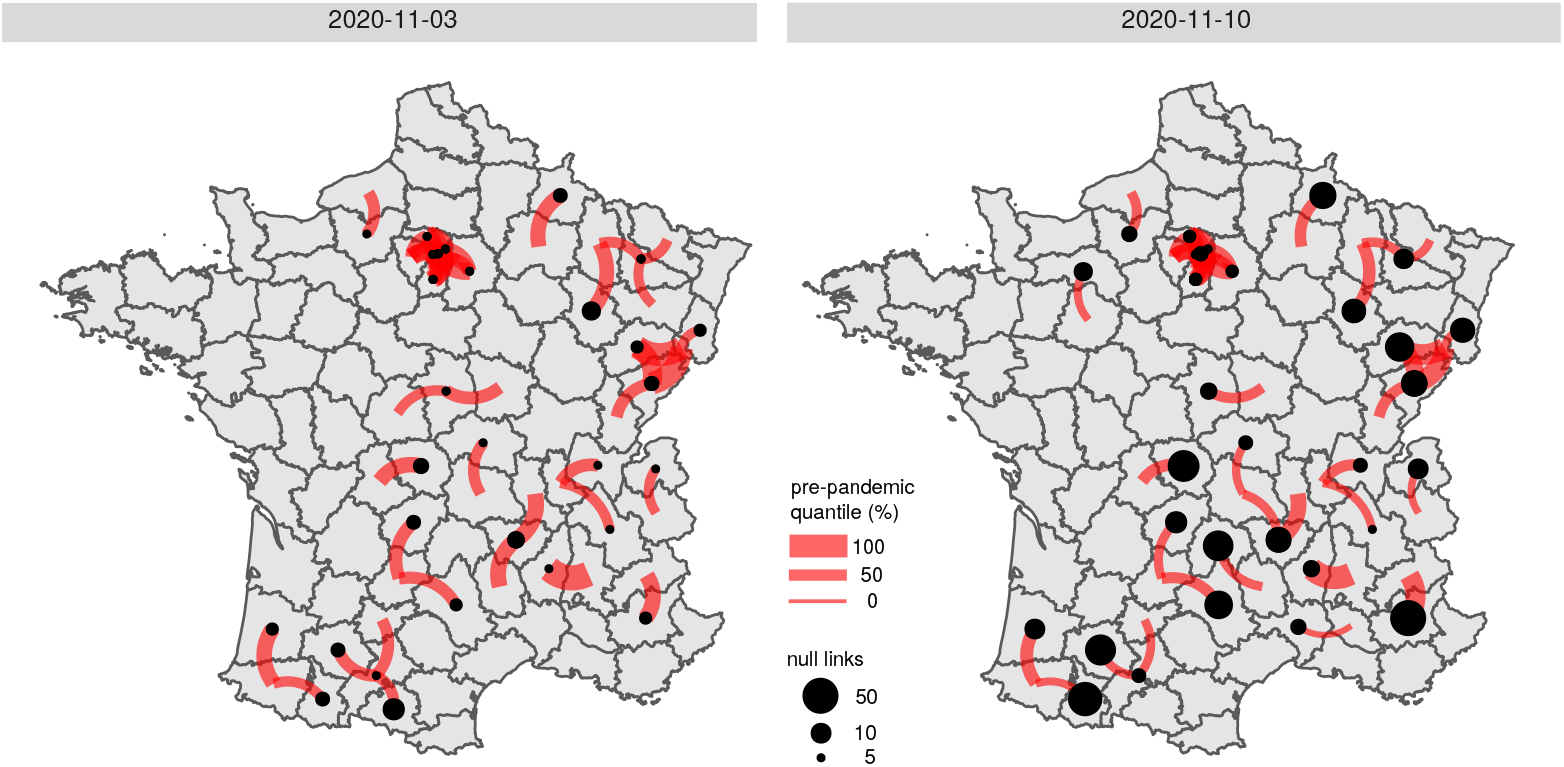
Fifty strongest between-département colocation links and null links from those départements in France, at weeks November 3rd (left panel) and November 10th, 2020 (right panel). The thickness of the red edges refers to the colocation weights in terms of pre-pandemic quantiles, the size of the black dots shows the number of missing links towards other départements in the particular week.

Correlation plots (Fig S1) across all recorded weeks indicated a strong positive correlation between network strength, curvature and Google Mobility (retail, grocery, parks, transit), but negative correlations between these quantities and null links. Interestingly, within-département colocation was only weakly correlated with Google Mobility data, supporting the hypothesis that colocation data could yield signals qualitatively different from more classical individual movement data.

### 3.2. Predicting national-level hospital incidence

The time series analysis showed that predictions made using only incidence data from past time points led to an average mean error of 1,632 hospitalisations per week and 238 deaths per week (regressor group “none” in Fig 3). Including a single exogenous regressor across all time points clearly reduced the prediction error by about 13% for hospital admissions and by 50% for hospital deaths (Fig 3). Quantiles from Facebook colocation data such as network strength, clustering or null links were among regressors that reduced prediction errors most. Google Mobility data related to recreation and residential movement also greatly improved predictions regarding mortality but not hospital admissions. Positive testing rate only appeared to improve predictions with two-week horizons. Finally, temperature did not belong the best predictors for hospital admissions or mortality.

**Figure 3:**
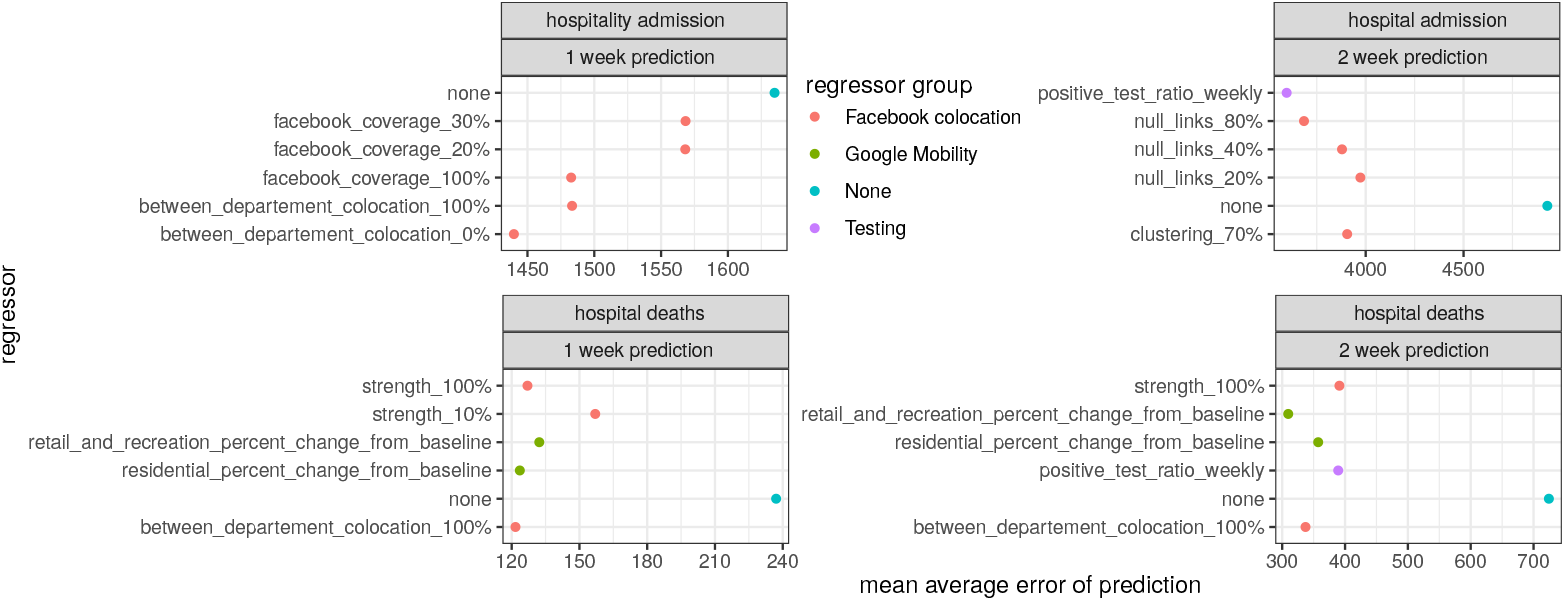
Top five predictive models for new hospital mortality (top) and admission (bottom). We show the models with the lowest mean average error using network analytics as regressors for one-week (left) and two-week (right) horizons. The error made in the model without exogeneous variable is shown in all panels. The colour code refers to the group of regressors used.

To further our understanding of temporal dynamics of hospital admissions, in particular at epidemic turning points, we determined for each week combinations of regressors that reduced the error most. For the one-week prediction horizon, we obtained almost perfect fits using regressor combinations mostly related to colocation networks (e.g. clustering and curvature quantiles) and, to a lesser extent, temperature (Fig 5). Although combining regressors improved the model fit at the two-week horizon compared to the model without exogenous regressors (the red curve), the model was not capable of reproducing the precision seen at the one-week prediction horizon. Similarly, combining regressors to predict hospital deaths resulted in almost perfect fits for the one-week horizon, and greatly improved fits for the two-week horizon, albeit slightly overestimating peak mortality (Fig 4).

**Figure 4:**
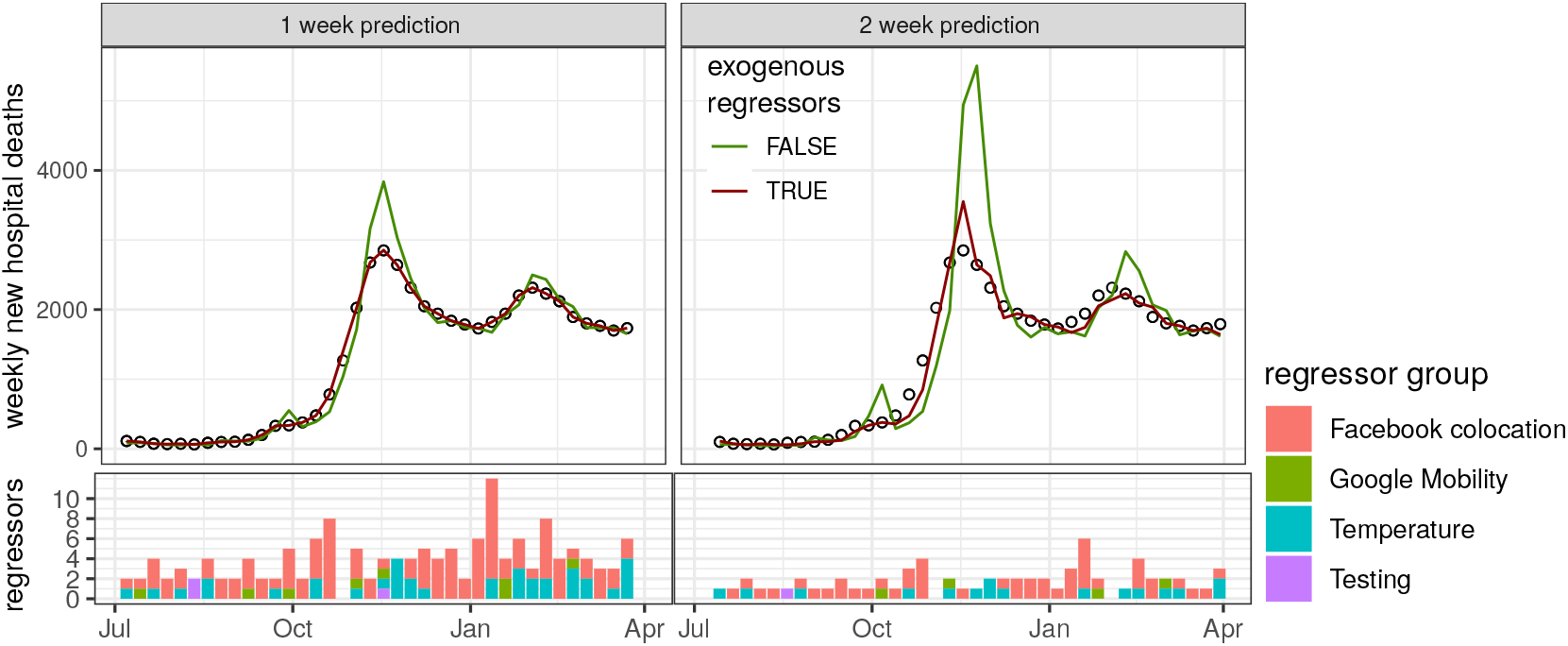
Model fits to hospital death incidence data (circles) with lowest mean average error among all models using combinations of regressors (red line) compared to auto-regressive models without exogenous regressors (green line). The bottom panels show the number of regressors used to obtain best fits, color-coded by the regressor group.

**Figure 5:**
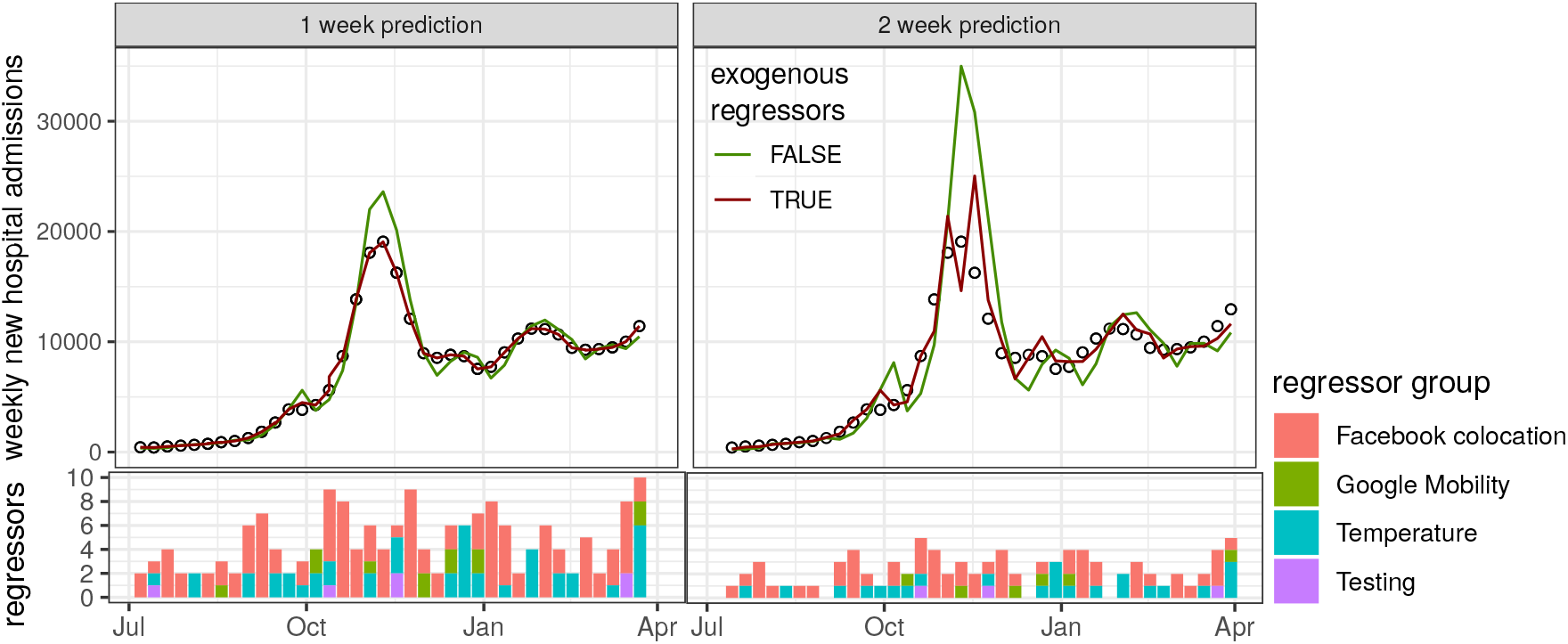
Model fits to hospital admission incidence data (circles) with lowest mean average error among all models using combinations of regressors (red line) compared to auto-regressive models without exogenous regressors (green line). The bottom panels show the number of regressors used to obtain best fits, color-coded by the regressor group.

### 3.3. Predicting département-level hospital incidence

Performing the same analysis at the département level revealed the importance of spatial structure. In particular for Paris region départements (Paris, Yvelines, Seine-Saint-Denis, Seine-et-Marne) and Nantes, the predictors that decreased the mean prediction error most when compared to models without regressors were those related to the incidence in highly colocalized départements (Fig 6). Conversely, for the metropolitan areas of Lyon, Bordeaux, and Marseilles, network descriptors (in red) such as curvature, null links, but also overall between-département colocation weights were among the best predictors of hospital admissions and deaths. This was also the case for two-week predictions of local hospital deaths, and to a lesser extent for two-week prediction of hospital admissions (Fig 7).

**Figure 6:**
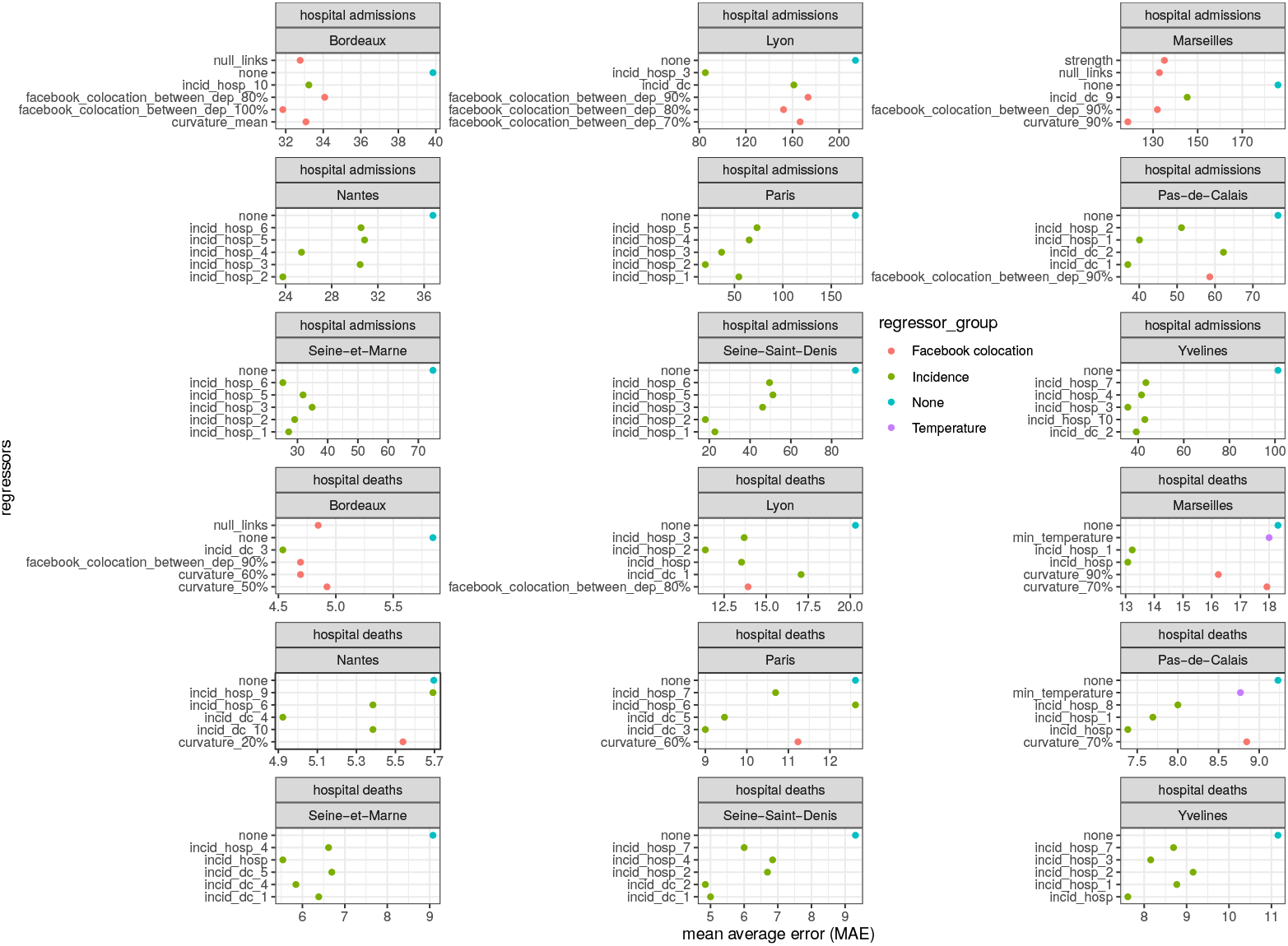
Top five one-week predictive models for new département-level hospital admissions and deaths with lowest mean average error using network analytics or incidence (‘dc’ for death and ‘hosp’ for hospital admission) of the top 10 colocated départements as regressors (numbers in subscripts). The colour code refers to the group of regressors used and the panels refer to the 9 most populated départements used in the analysis.

**Figure 7:**
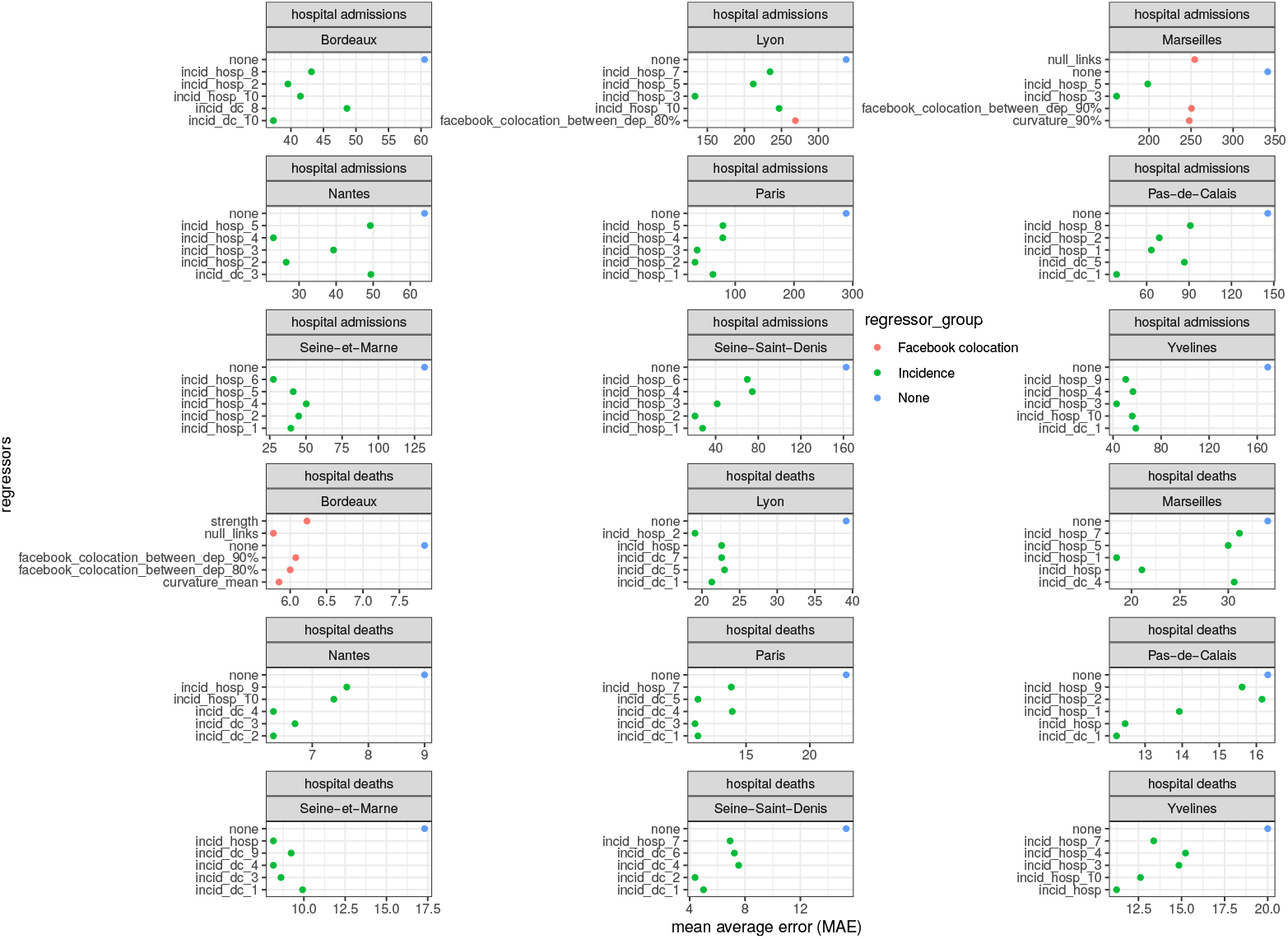
Top five two-week predictive models for new département-level hospital admissions and deaths with lowest mean average error using network analytics or incidence (‘dc’ for death and ‘hosp’ for hospital admission) of the top 10 colocated départements as regressors (numbers in subscripts). The colour code refers to the group of regressors used and the panels refer to the 9 most populated départements used in the analysis.

## 4. Discussion

Numerous studies have put forward the potential insights that can be gained from mobile phone usage data in order to understand the spread of infectious diseases. However, this data is usually analysed from an individual perspective, by following where the users are and how they move. The Facebook Inc. data differs in that it contains colocation data. We hypothesised that such data could be particularly suited to understand the transmission of respiratory infections, which involves close contact between individuals. To address this question, we developed an explicit network-based analysis to analyse relevant summary statistics regarding colocation data.

Our analysis indicates that human mobility and contact data improves time series prediction of French COVID-19 hospital admissions and deaths by up to 50%. Determining a posteriori the optimal combination of regressors shows that human contact network analytics augmented with temperature and positive testing rate data yields perfect fits at a one-week horizon. Although these combinations are not of predictive nature, they highlight the impact of global network properties, which are a proxy of the extent of human contact in contrast to human mobility alone. Spatial disparities in disease incidence have motivated sub-national policy decisions to manage the pandemic [25], and our sub-national time series analysis confirms the predictive power of spatial structure since incidence from colocalized administrative units improved most incidence predictions. Notably, we find that human contact network analytics such as curvature, null links, and network strength are prominent predictors for the metropolitan areas of Lyon, Bordeaux, and Marseilles. Further investigations using a meta-population point of view could yield additional insights.

Another asset of our study is that even though our two-week prediction horizon is relatively large compared to other studies, it remains quite robust for predictions of hospital deaths when compared to one-week predictions, even for a posteriori fits. This is also the slightly the case for hospital admissions. The fact that peak incidence was systematically overestimated points to the weakness of using only linear models, especially in situations where the susceptible populations are rapidly depleted.

Our results reveal the contrasting importance of temperature fluctuations, since temperature was not included in the top univariate regressors, but improved a posteriori fits when combined with other regressors. One possible explanation could be that national and sub-national policy changes over the time period considered led to human contact mobility regressors to be favored over temperature in terms of explanatory power.

Our study has several limitations. First, by focusing on the elaboration of contact network analytics we have utilized rather elementary statistical models. These have the advantage to be easy to interpret, but, given the number of features, more sophisticated machine learning techniques such as long short-term memory neural networks or random forests might could allow extracting more information from the data. Second, the interpretation of colocation data as a proxy for infectious contacts remains to be validated in the field, *e*.*g*. in community transmission studies [26]. The validation would also require to concurrently taking into account the importance of hospital catchment areas when recording disease incidence [27]. Furthermore, assortativity between users and user coverage might introduce biases that do not reflect the extent of human contacts pertaining to disease transmission.

Taken together, our time series analysis shows the potential of human contact network analytics to improve both predictions and a posteriori model fits of disease incidence as recorded during the COVID-19 pandemic in France. Combining network analytics with mechanistic models of disease transmission opens promising novel avenues for real-time disease control.

## Data Availability

Data and code will be made available through the zenodo repository.

https://www.facebook.com/geoinsights-portal

https://www.google.com/covid19/mobility

https://covid.ourworldindata.org

https://data.gouv.fr

## 5. Acknowledgements

We thank the Data for Good initiative by Facebook Inc. who provided the co-localization data through a data sharing agreement with the University of Montpellier.

The authors thank the CNRS, the IRD, and acknowledge the itrop HPC (South Green Platform) at IRD Montpellier for providing HPC resources that have contributed to the research results reported within this study (https://bioinfo.ird.fr/).

---

**Figure S1:**
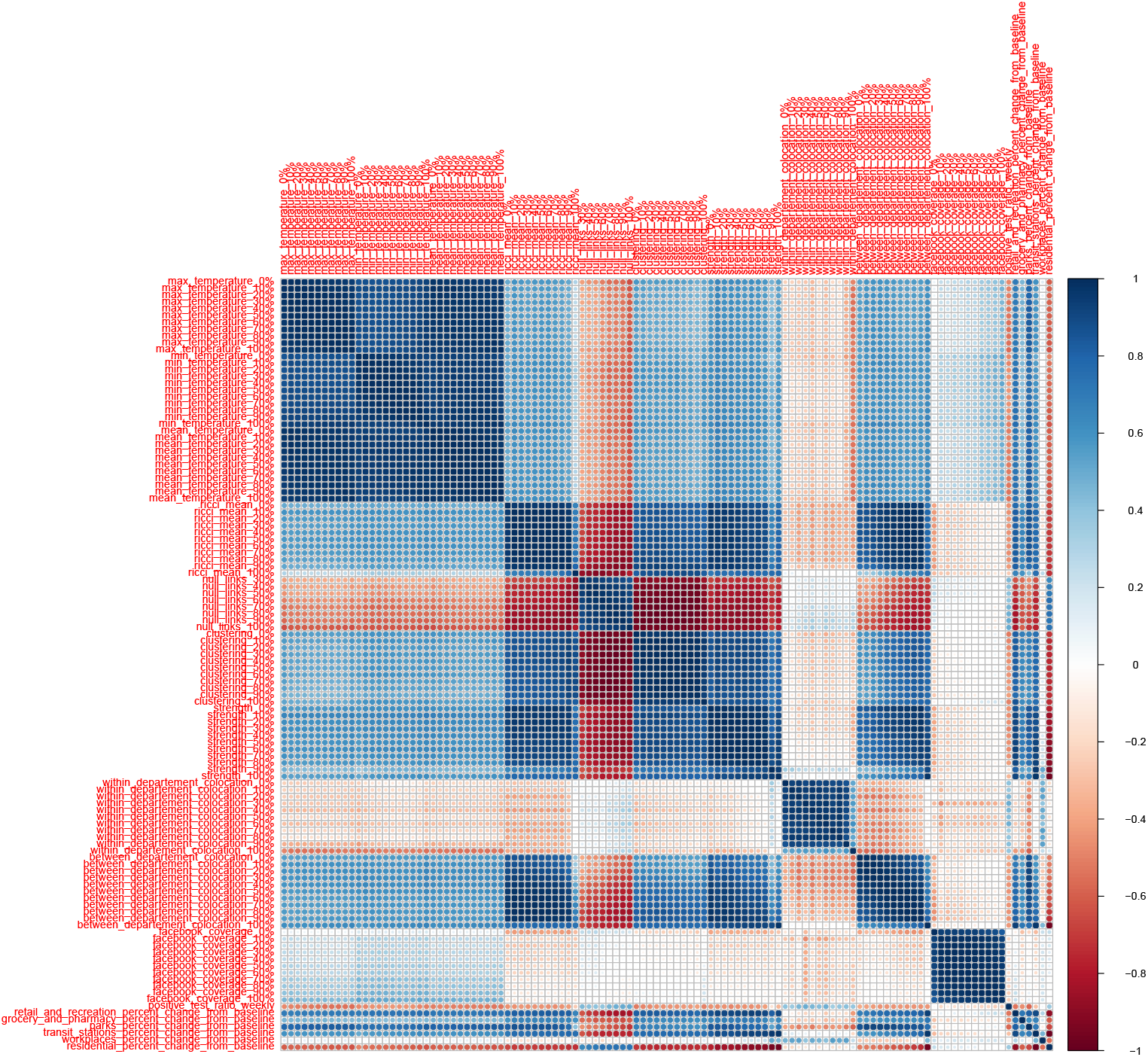
Correlation diagram of regressors.

## Notes

### Competing Interest Statement

The authors have declared no competing interest.

### Author Declarations

The source data for the study were openly available before the initiation of the study and can be accessed without application, or screening, or registration for access

